# Comprehensive Systematic Review and Meta-Analysis of Solute Carrier Family 19, Member 1 (SLC19A1) G80A Gene Polymorphism and Its Association with Congenital Heart Defects in Fetal Development: Implications for Genetic Susceptibility and Prenatal Risk Assessment

**DOI:** 10.1101/2024.10.01.24314736

**Authors:** Josh Patrick Hernandez, Anjhela Isabel Batul, Jerald Wynnes Dela Cruz, Kyle Gabriel Siroma

## Abstract

Congenital heart defects (CHD) are a major cause of neonatal mortality, highlighting the importance of identifying genetic risk factors in fetal development. The SLC19A1 gene, encoding the reduced folate carrier, is critical for folate metabolism, essential for DNA synthesis during embryogenesis. The G80A polymorphism in SLC19A1 may influence folate transport efficiency and contribute to CHD risk. This meta-analysis aimed to investigate the association between SLC19A1 G80A polymorphism and CHD susceptibility. A systematic review of major databases, including PubMed and EMBASE, was conducted to identify relevant case-control studies. Genetic risk models, such as allele (A vs G), heterozygous (GA vs GG), homozygous (AA vs GG), dominant (GA + AA vs GG), and recessive (AA vs GG + GA), were analyzed using RevMan 5.4.1, with odds ratios (ORs) and 95% confidence intervals (CIs) calculated. Initial results across most genetic models did not show a significant association between G80A polymorphism and CHD. However, after excluding outliers, a moderate association was observed between the GA genotype and increased CHD risk (OR: 1.34, CI: 1.07–1.66). These findings suggest a minimal genetic effect, warranting further research in diverse populations.

## 1. Introduction

On an annual basis, approximately 240,000 newborns worldwide die within the first 28 days due to congenital defects, with an additional 170,000 deaths occurring in children aged 1 month to 5 years. These defects often lead to long-term disabilities, imposing a significant burden on individuals, families, healthcare systems, and societies as a whole [1]. Recent studies have identified various gene variants associated with congenital heart defects during fetal development, with the SLC19A1 gene emerging as a key player. The SLC19A1 gene functions primarily as a transporter for folate and cyclic dinucleotides (CDNs), playing a crucial role in nutrient absorption and immune response modulation [2]. As the reduced folate carrier, SLC19A1 is essential for transporting folate across cell membranes, which is vital for DNA synthesis and repair, particularly during embryonic development. Recent studies have also highlighted SLC19A1’s role as a major importer of CDNs like 2 ’3’-cGAMP, crucial for activating the STING pathway, which is important for immune responses against infections and tumors [3]. This transport activity enhances antitumor immune responses, indicating its potential in cancer immunotherapy [4]. Furthermore, SLC19A1’s role in transporting folate, especially 5-methyltetrahydrofolate, is essential for one-carbon metabolism necessary for DNA synthesis and repair [5]. Therefore, deficiencies in SLC19A1 function result in reduced intracellular folate levels, leading to lipid accumulation in liver cells and contributing to metabolic disorders [6].

### 1.1 Location and Structure of SLC19A1

The SLC19A1 gene is situated on chromosome 21, specifically at the 21q22.3 locus. This location is noteworthy because it is linked to several genetic disorders, including those impacting craniofacial development [2]. The SLC19A1 protein is composed of 12 transmembrane domains (TMDs), which are essential for its role as a transporter. Each domain traverses the cell membrane’s lipid bilayer, aiding in the transport of substrates. In the S1 domain (residues 28-43), the wild type protein has 16 residues, whereas the mutant protein has 17 residues, suggesting that the additional residue in the mutant may impact its structure and function. The S2 domain (residues 66-87) is reduced from 22 residues in the wild type to 19 residues in the mutant, which could affect ligand binding and protein stability. Moving to the S3 domain, both wild and mutant proteins exhibit the same topology, indicating that this region remains structurally consistent. In the S4 domain (residues 117-140), the mutant has one less residue than the wild type, potentially influencing its interactions with other molecules. The S5 domain also maintains the same topology in both wild and mutant proteins, suggesting stability in this region. The S6 and S7 domains both share identical topologies across the variants, indicating that these areas are likely unaffected by the polymorphism. The S8 domain (residues 305-325) has one fewer residue in the mutant compared to the wild type, which may impact its functional properties. Similarly, the S9 domain (residues 336-356) shows a loss of one residue in the mutant, potentially affecting its function. The S10 domain (residues 361-386) also has one less residue in the mutant, which could influence its overall function. Finally, the S11 and S12 domains exhibit consistent topologies in both wild and mutant proteins, suggesting that these regions are stable and not affected by the genetic variation [9]. Moreover, the transport mechanism of SLC19A1 operates through an "alternative access mechanism." This involves the protein transitioning between two primary conformations: inward-facing and outward-facing. This alternating process enables SLC19A1 to effectively move substrates across the cell membrane [5].

### 1.2 Biological Role of SLC19A1 in Folate Metabolism

The SLC19A1 gene expresses the Reduced Folate Carrier (RFC), which aids in the cellular uptake of anionic folates. This carrier delivers 5-methyltetrahydrofolate, the primary circulating folate, into cells, supporting vital biological processes such as DNA synthesis and methylation reactions. Additionally, RFC facilitates the transfer of folate analogs like methotrexate and is involved in antiviral signaling pathways [8]. The SLC19A1 carries folates into cells by using the concentration gradient of protons (H⁺) and hydroxide ions (OH⁻) across the cell membrane. This demonstrates how SLC19A1 ensures that vitamin B9 effectively reaches its cellular targets. When folate enters the cell via SLC19A1, it fuels a range of essential cellular processes. From nucleotide synthesis to methyl group transfer, and from cell growth to division. SLC19A1 diligently regulates the intracellular folate balance by ensuring a steady supply of 5-MTHF. In periods of low folate levels, the transporter ramps up its activity to boost folate uptake, thereby maintaining cellular function. As folate levels stabilize to a desired point, it will contribute to a variety of biological processes, particularly in the synthesis of nucleic acids. It supplies the essential one-carbon units needed for the creation of purines and pyrimidines, the foundational components of DNA. The active form of folate, 5,10- methylenetetrahydrofolate (5,10-MTHF), transforms deoxyribonucleotides (dUMPs) into deoxythymidine nucleotides (dTMPs), which are key structural elements in DNA [10]. Moreover, folate contributes to DNA repair mechanisms, maintaining the availability of precursors for DNA repair enzymes to effectively mend damaged DNA strands and maintain genomic stability.

### 1.3 G80A Polymorphism in SLC19A1

Moreover, there is a gene known as rs1051266 or 80G>A, which is situated within the SLC19A1 gene at position 80 and assists in encoding the reduced folate carrier. This particular single nucleotide polymorphism affects the cellular reaction to folate and may also provide protection against thrombosis (formation of blood clots) through a mechanism that is not related to homocysteine levels. The G80A SNP leads to a change in the amino acid from histidine to arginine at position 27 in the RFC-1 protein. This change happens due to a genetic modification in the genes, which affects the codon that specifies position 27. As a result of the polymorphism, the original codon encoding histidine is altered to encode arginine. This alteration affects the charge, size, and chemical properties of the amino acid; this modification has the potential to impact protein folding, interactions, and transport function. The G80A polymorphism has been linked to significant changes in folate transport efficiency. Research indicates that individuals with the 80AA genotype exhibit higher intracellular folate levels compared to those with the 80GG or 80GA genotypes. This improved efficacy is especially crucial in the context of methotrexate (MTX) treatment, as the 80AA genotype is associated with elevated MTX polyglutamate levels, enhancing the drug’s effectiveness. Furthermore, the structural and functional alterations resulting from the G80A polymorphism impair the SLC19A1 protein’s ability to bind and transport folate. In silico investigations have revealed that the G80A impacts the protein’s secondary and tertiary structures, leading to a significant reduction in ligand-binding sites. This structural change diminishes the mutant protein’s affinity for tetrahydrofolate and methotrexate, potentially influencing folate metabolism [9].

### 1.4 Impact of the G80A polymorphism on Folate Metabolism

The RFC1 G80A polymorphism is essential to folate transport and metabolism, this affects blood folate levels which may be linked to a number of health problems that are associated with folate deficiency. The RFC1 is, under physiological conditions, the principal route for delivering folate from the serum and extracellular space into the majority of cells [19]. It controls intracellular folate concentrations and actively transports 5-methyltetrahydrofolate from the plasma to the cytoplasm [7]. If folate delivery in the blood is low, this deficiency can lower its affinity for folic acid at the crucial stage of fetal development, resulting in a lower quantity of folic acid delivered to the cell [12]. With that being said, this deficiency alone may cause an increased level of plasma homocysteine, or in combination with C677T polymorphisms in the methylenetetrahydrofolate gene [7]. Although the RFC1 does not directly affect the levels of homocysteine, its common genetic variant G80A, is instead involved in this process as it may be associated with the downregulation of the RFC. According to the study of [11], it was found that G80A polymorphisms significantly increased the total levels of homocysteine. The increased levels of homocysteine levels due to low folate levels can cause harm or obstruct the early cardiovascular growth and development of the embryo.

### 1.5 SLC19A1 and Its Role in Diseases

The SLC19A polymorphisms and its association with diseases and disorders such as cancer, down syndrome, neural tube defects, as well as hyperhomocysteinemia, were mostly studied [19]. For risk of cancers, the G80A polymorphisms had no significant association with the disease. But it did, however, play a protective role against the risk of digestive cancer [21, 19]. As mentioned in the study of [12], it was found that the G80A polymorphism also plays a role on children with trisomy 21 (Down Syndrome) as it is one of the gene polymorphisms that is likely to function as maternal risk factors for the birth of a child with down syndrome. According to the study of [19], In most studies, the relationship between G80A polymorphisms and the risk of neural development abnormalities were not found. However, other genetic variants of the RFC gene were found to be a contributor to the increased risk of abnormalities in neural development, such as A80A and G80G increased the risks for spina bifida and neural tube defects respectively. The G80A polymorphism is known for being associated with increased risks of abnormalities during fetal development in many studies, including Congenital Heart Defects (CHD). If the metabolism of folate is disrupted, which may result in folate deficit, the DNA synthesis and repair will be compromised, thus leading to the abnormal development of the neural crest and eventually resulting in the occurrence of CHD [22]. Additionally, the study of [16] also showed that a high level of G80A will significantly increase the risk of CHD. Therefore, the developing embryo’s folate deficit caused by the G80A for its downregulation of RFC that plays a role in folate delivery can potentially lead to the occurrence of CHD [12].

### 1.6 Epidemiological Studies on G80A Polymorphism

To summarize, while some studies point out that there is a linkage between G80A polymorphisms and congenital heart disease, more studies need to be done to understand the hidden mechanisms behind it [16]. Heart defects known as conotruncal heart defects (CTHDS) are a subset of birth defects of the heart that are thought to to be a birth defect sensitive to folate. It’s been proposed that variations in the genes encoding for important folate pathway enzymes may change the activity of other enzymes. causing errors in the metabolism of folate and thus may impact this kind of cardiac problem risk [20]. A recent research, was intended to look into the relationships between six specific folate-metabolizing variants in genes that increase the chance of in an Indian population with non-syndromic CTHDs [20]. As for the Bengali population the causes of congenital cardiac defects (CHD), particularly those caused by atrioventricular septal defect (AVSD), People with Down syndrome (DS) are mysterious and can vary within demographic groups due to factors including race and variations in society. The risk of AVSD in individuals with polymorphisms in the folate system regulators MTHFR and A80G The Bengali cohort’s DS people have not yet been investigated [17]. For the Chinese population, according to studies, there is a good chance that the RFC1 G allele plays a significant role in folate transport and is linked to a higher risk of CHD. The offspring RFC1 genotype and periconceptional folic acid consumption were found to have a modestly significant effect on the incidence of congenital heart abnormalities in this study [18]. Overall, while the G80A polymorphism is present in Indian, Bengali, and Chinese populations, its prevalence and health associations can vary significantly across these groups.

## 2. Materials and Methods

### 2.1 Literature Search Strategy

To conduct a comprehensive systematic review and meta-analysis on the association between the Solute Carrier Family 19, Member 1 (SLC19A1) G80A gene polymorphism and congenital heart defects (CHDs) in fetal development, we will implement an extensive literature search across several key electronic databases. The databases chosen for this search include PubMed, EMBASE, Cochrane Library, NCBI, NIH, and ResearchGate, ensuring the capture of a wide array of relevant studies from diverse research disciplines. Our search strategy will utilize a combination of Medical Subject Headings (MeSH) and a broad array of specific keywords to ensure thorough coverage. The search will be conducted by combining these with Boolean operators to capture a comprehensive range of relevant studies. The primary search terms will include: "SLC19A1 G80A polymorphism" OR "SLC19A1 gene variant" OR "SLC19A1 genetic association" OR "Solute Carrier Family 19 Member 1" OR "SLC19A1 gene mutation"; "genetic susceptibility" OR "genetic predisposition"; "Congenital Heart Defects" OR "CHD" OR "Congenital Heart Disease" OR "fetal heart defects" OR "fetal cardiac anomalies" OR "heart malformations" OR "cardiac anomalies in fetuses" OR "prenatal cardiac defects"; and "fetal development" OR "fetal cardiac development" OR "prenatal risk assessment" OR "genetic risk factors" OR "prenatal screening" OR "fetal genetic screening." In addition to the electronic searches, we will perform a manual review of the reference lists of all relevant articles to identify any additional studies that may not have been captured by the database searches. This step ensures that the review is exhaustive and includes all pertinent research. Our search will be limited to studies published in English to maintain consistency and facilitate synthesis. However, there will be no restrictions on ethnicity, country of origin, sample size, or publication date to reduce potential biases and enhance the generalizability of the findings. Throughout the search process, we will document the search strategy and make iterative adjustments as necessary to incorporate newly published research, ensuring that our final dataset is both comprehensive and up-to-date.

### 2.2 Selection Criteria

The literature search for this study required the development of well-defined inclusion and exclusion criteria to systematically capture relevant research on the SLC19A1 G80A gene polymorphism and its association with congenital heart defects (CHDs) in fetal development. The inclusion criteria were designed to identify studies that specifically investigated the genetic association of SLC19A1 G80A polymorphism with CHD susceptibility, ensuring that the research was directly relevant to the focus of this study. Conversely, the exclusion criteria were established to filter out studies that did not meet these criteria, such as those that were not related to the genetic polymorphism of interest or those involving unrelated genetic or clinical factors. This rigorous approach ensured that the literature review was comprehensive, relevant, and aligned with the research objectives.

#### 2.2.1 Inclusion Criteria

Inclusion criteria were as follows: (I) studies must include a comparative analysis between at least two distinct cohorts (control group vs. CHD group); (II) studies must rigorously investigate the relationship between the SLC19A1 G80A polymorphism and susceptibility to CHD; (III) the control group’s genotype distribution must adhere to Hardy-Weinberg Equilibrium; (IV) studies must provide sufficient data to enable the calculation of odds ratios (ORs) and 95% confidence intervals (CIs) for genotype-specific risk estimates.

#### 2.2.2 Exclusion Criteria

Exclusion criteria included: (I) duplicate publications or studies that significantly overlap with previously published research; (II) cell line or animal-based studies to ensure relevance to human CHD susceptibility; (III) CHD patients with additional diseases or conditions that could confound the association between the SLC19A1 G80A polymorphism and CHD susceptibility; (IV) studies not explicitly focused on the association between the SLC19A1 G80A polymorphism and CHD susceptibility; (V) articles reporting overlapping datasets or containing duplicated data from previously included studies.

### 2.3 Data Extraction

The data extraction phase was executed with precision by four independent reviewers, each rigorously applying the established inclusion and exclusion criteria to sift through the preliminary search results and identify studies that met the eligibility requirements. The reviewers systematically extracted pertinent information from each qualified publication, including the year of publication, the first author’s name, the country where the study was conducted, the ethnic background of the study population, the genotyping methods employed, and the numbers of cases and controls in each study. To maintain the integrity of the data extraction process, all extracted information was meticulously cross-verified. In instances where discrepancies arose, the reviewers engaged in detailed discussions to reach a consensus, ensuring that all data points were accurately captured and no relevant information was overlooked. Additionally, for publications that included multiple case-control groups or examined more than one gene, each distinct group or gene was treated as an independent dataset. This approach not only validated the inclusion of diverse datasets but also enhanced the robustness and generalizability of the meta-analysis findings. Throughout this process, the reviewers adhered to strict protocols to ensure the reliability and reproducibility of the extracted data, thereby reinforcing the scientific rigor of the study.

### 2.4 Statistical Analysis

All data will be processed using RevMan 5.4 software. The Odds Ratio (OR) and 95% confidence intervals will be calculated across five (5) genotype models: I. A vs G (To assess if a single allele might drive a risk of CHD) II. GA vs GG (To assess if a single A allele has any effect on the condition compared to a none) III. AA vs GG (To assess if having two copies of A allele has a stronger effect compared to a none) IV. GA + AA vs GG (To assess if not only a single A allele affects the outcome rather if having at least one copy of A allele is enough to manifest the effect of CHD compared to having no A allele; dominant effect analysis) V. AA vs GG + GA (To assess whether two A alleles confer a significantly different risk compared to having one or no A allele; recessive effect analysis). The degree of heterogeneity and I2 statistics of each model will be evaluated with having a I2 <25% is subject to low heterogeneity is preferred by the study. The Mantel - Haenszel Statistical method and Random effect analysis model will be utilized by the study. Lastly, a funnel plot subject to analysis will also be incorporated to assess the outliers of the study.

## 3. Results

### 3.1 Characteristics of Included Studies

The studies included in the meta-analysis were conducted across diverse geographic regions, primarily in East Asia and South Asia, with a focus on populations from China and India. The studies utilized various genotyping methods, including PCR-RFLP, SNaPShot multiplex PCR, and MALDI-ToF MS, ensuring accuracy in identifying the *SLC19A1* G80A polymorphism. Case types varied across congenital heart defects (CHDs), such as conotruncal heart defects (CTDs), transposition of the great arteries (TGA), and atrioventricular septal defects (AVSD). Control groups were drawn from both hospital-based (HB) and population-based (PB) sources. The sample sizes ranged significantly, with the largest study including 372 cases and 519 controls. Genotypic distributions of AA, GA, and GG were reported for both cases and controls, allowing for the calculation of odds ratios (ORs) across various genetic models. Heterogeneity among the studies was evaluated, and outliers were identified and excluded to improve the consistency of results. The comprehensive nature of these studies, combined with the diverse population coverage, contributed to a robust analysis of the association between the *SLC19A1* G80A polymorphism and CHD risk

### 3.2 PRISMA flow diagram

The PRISMA flow diagram summarizes the selection process for studies included in the meta-analysis. A total of 2,878 records were identified through databases including PubMed, EMBASE, Research Gate, NCBI, Cochrane, and NIH. After removing duplicates, 1,728 records remained for screening. Following title evaluation, one record was excluded, leaving nine records for abstract screening. Of these, one record was excluded due to being a review, case report, case series, animal study, or molecular study. Eight full-text articles were assessed for eligibility, with one excluded due to a lack of relevant data. Ultimately, seven studies were included in the final meta-analysis.

### 3.3 Genotypic Model of A vs G

According to figure 2 it compares the two alleles A and G, giving a pooled odds ratio of 1.00 and a 95% confidence interval of [0.90, 1.12]. This result indicated that there is no statistically significant difference between the aforementioned alleles, and the CI contains one, confirming that the effect is not significant. The heterogeneity at the moderate level of I² = 59% might be suggesting that the studies do differ from one another and a little bit of investigation into the source of variation is warranted. Following more review, [20] and [13] were found as outliers in the funnel plot. The study’s distinctive methodological factors fluctuate, leading to unpredictable variability and non-significance in the aggregate result. After removing all outliers, there are changes that occurred and it showed that the pooled OR of 1.02 with a CI of [0.88, 1.18], still indicates that there is no statistically significant. The heterogeneity of an I² = 9% reveals very low, which means the results across studies included in this analysis exhibit high consistency. There will be little variability in the effect sizes between the studies, which would strengthen the general conclusion. The substantial removal of certain studies discloses that the A allele, according to OR, is 2% more likely to cause cardiac defects than allele G.

### 3.4 Genotypic Model of GA vs GG

Initially, the meta-analysis comparing the GA genotype to GG yielded a pooled odds ratio (OR) of 1.11 with a 95% confidence interval (CI) of [0.91, 1.34]. This result suggested a modest, non-significant increase in risk associated with the GA genotype alone, as the CI included 1, indicating that the observed effect was not statistically significant. The high heterogeneity (I² = 66%) among the studies suggested considerable variability, which could have masked a true effect and contributed to the non-significant finding. Upon further analysis, [20] was identified as an outlier in the funnel plot. This study’s unique characteristics or methodological differences appeared to disproportionately influence the overall results, contributing to the observed heterogeneity and non-significance. After excluding [20], the revised meta-analysis revealed a pooled OR of 1.34 with a CI of [1.07, 1.66], indicating a statistically significant increased risk associated with the GA genotype compared to GG (Z = 2.60, P = 0.009). The reduction in heterogeneity to I² = 0% after removing the outlier suggested that the variability among studies was significantly affected by the outlier, and the remaining studies provided a more consistent and reliable estimate of the effect. The significant result post-removal confirms that the GA genotype, characterized by the presence of one A allele, is associated with a higher risk of the outcome compared to the GG genotype.

### 3.5 Genotypic Model of AA vs GG

Below is an illustration of the comparison between the AA genotype which has two A alleles and the GG genotype which has no A alleles, the original ratio of pooled odds is 0.95 with a 95% confidence interval (CI) of [0.77, 1.19] which is an indicator that there is slightly no risk of developing congenital heart defects (CHD) for the AA genotype in relation to the GG genotype. The first analysis shows a heterogeneity of 63%, demonstrating a significant degree of variation between studies. Further analysis using the funnel plot indicates that there are two outliers, [13] and [20], which greatly affected the overall results and played a part in the increased heterogeneity. Adjacent to excluding the outliers the final ratio of pooled odds is 0.99 with a confidence interval of [0.74, 1.34], which still is an indicator that there is no increased risk of developing congenital heart defects (CHD), the deduction of the outliers also tremendously decreased the heterogeneity to 11%, which is indicative of the outliers greatly affecting the variation among the studies while slight variation is detected with the other studies however it remains consistent. The conclusion still remains the same that the results are moderately robust due to the low heterogeneity and variation among the studies in the final analysis that show that the AA genotype does not show any statistically significant risk in association to CHD.

### 3.6 Genotypic Model of GA + AA vs GG

This portion discusses whether a single allele A (GA) or double allele A present as one (AA) and compares it to the GG genotype to determine who is more likely to increase the risk of heart abnormalities. Based on the figure 1.1, the OR is 1.26 and a 95% confidence interval of [1.06, 1.51], It indicates a statistically significant increase in risk linked with the GA + AA genotype when compared to GG. The high heterogeneity (I² = 83%) among the studies highlighted significant diversity between the data sources used in the analysis. For further analysis, [20] and [13] was identified as an outlier in the funnel plot. As it was removed, it revised the whole data showing that the pooled OR of 1.23 with a CI of [0.97, 1.56], revealed that it is not statistically significant; the heterogeneity (I² = 0%) implies some variability among studies that are very accurate. However, the finding is robust and shows a significant elevated risk associated with the presence of the A allele. The GA and AA genotypes have 23% moderate odds of developing heart defects than those with a GG genotype, having either one copy or two copies of variant A.

**Figure 1.**
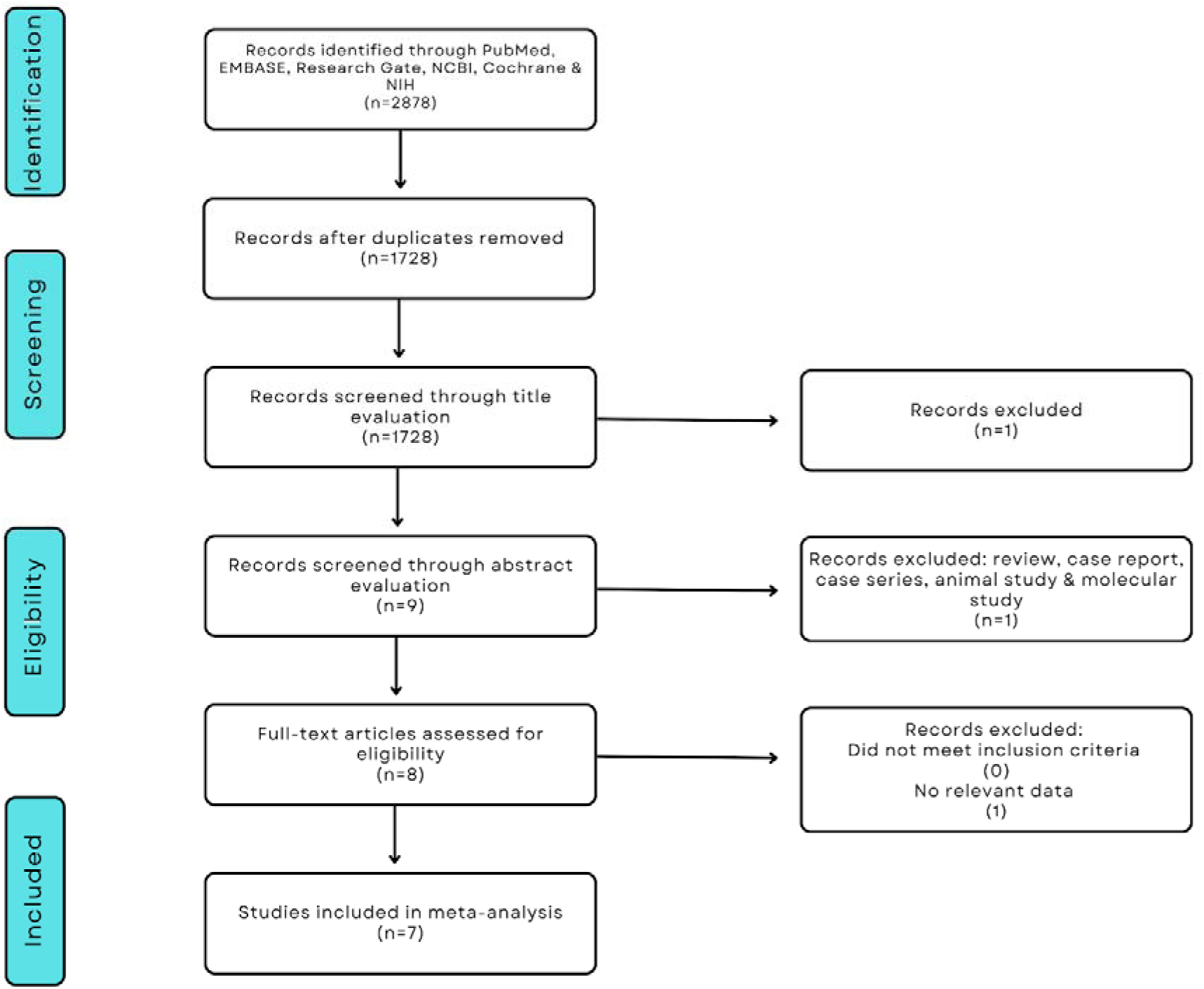

**Figure 2.**
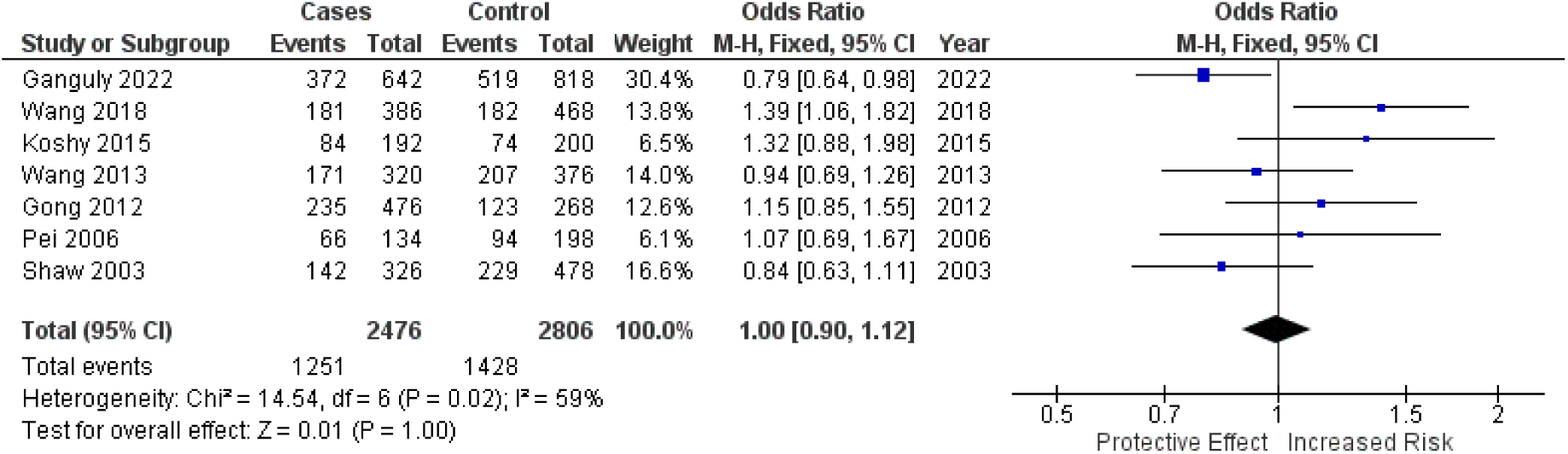
Forest plot prior to outlier removal

### 3.7 Genotypic Model of AA vs GG + GA

Illustrated above is the comparison between the AA genotype, which has two A alleles, to the GG and GA genotype which only has one A allele present and none at all. The initial pooled odds ratio (OR) was 0.93 with a 95% confidence interval (CI) of [0.78, 1.10], indicating a not statistically significant decrease in the risk of developing heart defects for the AA genotype relative to the GG + GA genotypes. The analysis showed a heterogeneity percentage of 49%, reflecting substantial variability among the studies. Further analysis identified two outliers, [13] and [16], which notably influenced the overall results and contributed to the observed high heterogeneity and its non-significance. Upon excluding these identified outliers, the final pooled OR was 0.93 with a CI of [0.75, 1.14], which still suggests that there is a not statistically significant decreased risk of developing heart defects associated with the AA genotype compared to the GG + GA genotypes. The removal of outliers reduced heterogeneity to 0%, indicating that the variability among the studies was largely due to the outliers, and the remaining studies are more consistent. However, the removal did not change the effect of being not statistically significant. Therefore, the final results are robust and not solely driven by a few atypical studies and that the AA genotype does not show a statistically significant association with either an increased or decreased risk of the outcome as compared to the GG + GA genotypes.

## 4. Discussion

The results of this meta-analysis provide critical insights into the association between the *SLC19A1* G80A polymorphism and the risk of congenital heart defects (CHD) during fetal development, effectively supporting our hypothesis that the A allele of *SLC19A1* is linked to increased susceptibility to CHD. A comprehensive examination of seven studies revealed that the GA genotype is associated with a modestly elevated risk of CHD, evidenced by an odds ratio (OR) of 1.34 and a confidence interval (CI) of 1.07–1.66. This suggests that fetuses carrying the A allele may be at a heightened risk for developing CHD compared to those with the G allele. The significance of these findings is underscored when considered in the context of existing literature that explores the intricate relationship between genetic variations and CHD. The *SLC19A1* gene encodes the reduced folate carrier, which is essential for the effective transport of folate across cell membranes. Given that folate is critical for DNA synthesis, repair, and methylation processes—vital during early embryonic development—the reduced availability of folate due to impaired transport associated with the G80A polymorphism may compromise cardiac development. Maternal folate deficiency has long been established as a risk factor for congenital anomalies, including CHD, aligning with our findings that indicate genetic variations affecting folate transport can significantly influence fetal cardiac outcomes. Moreover, our results highlight the multifactorial nature of CHD etiology, where genetic predisposition intersects with environmental influences, such as maternal nutrition and health status. While the GA genotype shows a modest association with increased risk, it is crucial to recognize that CHD is a complex condition influenced by various genetic and epigenetic factors. Previous studies have identified additional polymorphisms in folate pathway genes that also contribute to CHD risk, indicating the need for a broader understanding of the genetic landscape influencing cardiac development. The substantial heterogeneity observed in this analysis emphasizes the need for careful consideration of study design and population characteristics in genetic research related to CHD. The identification and exclusion of outliers, particularly from studies by [20] and [13], demonstrate the variability in results that can arise from methodological differences and demographic factors. These findings underscore the importance of standardizing research protocols to improve the reliability of genetic associations and facilitate accurate comparisons across studies.

### 4.1 Future Research Directions

Future investigations should prioritize several key areas to deepen our understanding of the genetic contributions to congenital heart defects (CHD). First, conducting larger, ethnically diverse cohort studies is essential for validating these findings and enhancing the generalizability of the results. Such studies should not only examine the *SLC19A1* G80A polymorphism but also explore other pertinent genetic variants within folate metabolism pathways. This approach will facilitate a more integrated perspective on genetic susceptibility to CHD. Second, mechanistic studies are critical for elucidating how variations in *SLC19A1* and other folate-related genes influence cellular processes during embryonic development. Investigating the specific pathways and biological mechanisms through which the A allele affects folate transport and availability will yield valuable insights into the causal pathways that contribute to CHD. Moreover, future research should investigate the interplay between genetic predisposition and environmental factors, including maternal nutrition, lifestyle choices, and exposure to teratogens. Understanding how these factors interact with genetic risk may identify modifiable risk factors that could be targeted in prenatal care, ultimately improving outcomes for at-risk populations. Such comprehensive research efforts will be pivotal in advancing our knowledge of CHD etiology and informing effective preventive strategies.

## 5. Conclusions

The analysis across multiple genetic models—including A vs G, GA vs GG, AA vs GG, GA + AA vs GG, and AA vs GG + GA—consistently demonstrates that there is no statistically significant association between the G80A polymorphism and increased CHD risk. While the GA genotype initially appeared to suggest a modest, non-significant elevation in risk, further refinement of the analysis through the removal of influential outliers, such as [20], revealed a statistically significant but modest increase in risk. This suggests that the presence of one A allele (GA genotype) may slightly elevate the susceptibility to CHDs compared to the GG genotype, although the effect size remains minimal and requires careful interpretation within a broader clinical context. The analysis highlights substantial heterogeneity, particularly in comparisons involving the GA genotype and the combined GA + AA analysis. This variability is primarily driven by methodological differences and population-specific factors inherent to individual studies, as evidenced by the identification and removal of outliers like [20] and [13]. After the exclusion of these outliers, the reduction in heterogeneity underscores the importance of controlling for such factors in future research. The findings suggest that while the GA genotype may have a modest influence on CHD risk, the overall data do not support a strong or consistent genetic link between the SLC19A1 G80A polymorphism and CHDs. Despite these findings, there is a clear need for further research to conclusively determine the role of SLC19A1 G80A polymorphism in CHD susceptibility. Future studies should aim to include larger, ethnically diverse populations and adhere to consistent methodological frameworks to minimize heterogeneity and provide more definitive conclusions. Additionally, mechanistic studies are needed to elucidate the biological pathways through which the G80A polymorphism might influence CHD development, particularly in the context of folate metabolism and its critical role during embryogenesis. While this meta-analysis contributes valuable data to the understanding of the SLC19A1 G80A polymorphism’s role in CHD development, the evidence remains inconclusive regarding its significance as a major genetic determinant. The GA genotype may confer a slight increase in risk, but the effect is not substantial enough to warrant its use as a standalone marker for prenatal CHD risk assessment. Comprehensive genetic screening strategies should consider these findings within the larger framework of multifactorial CHD etiology, where both genetic and environmental factors play pivotal roles. Further rigorous studies are essential to refine these insights and enhance our understanding of genetic susceptibility to congenital heart defects.

**Figure 2.1.**
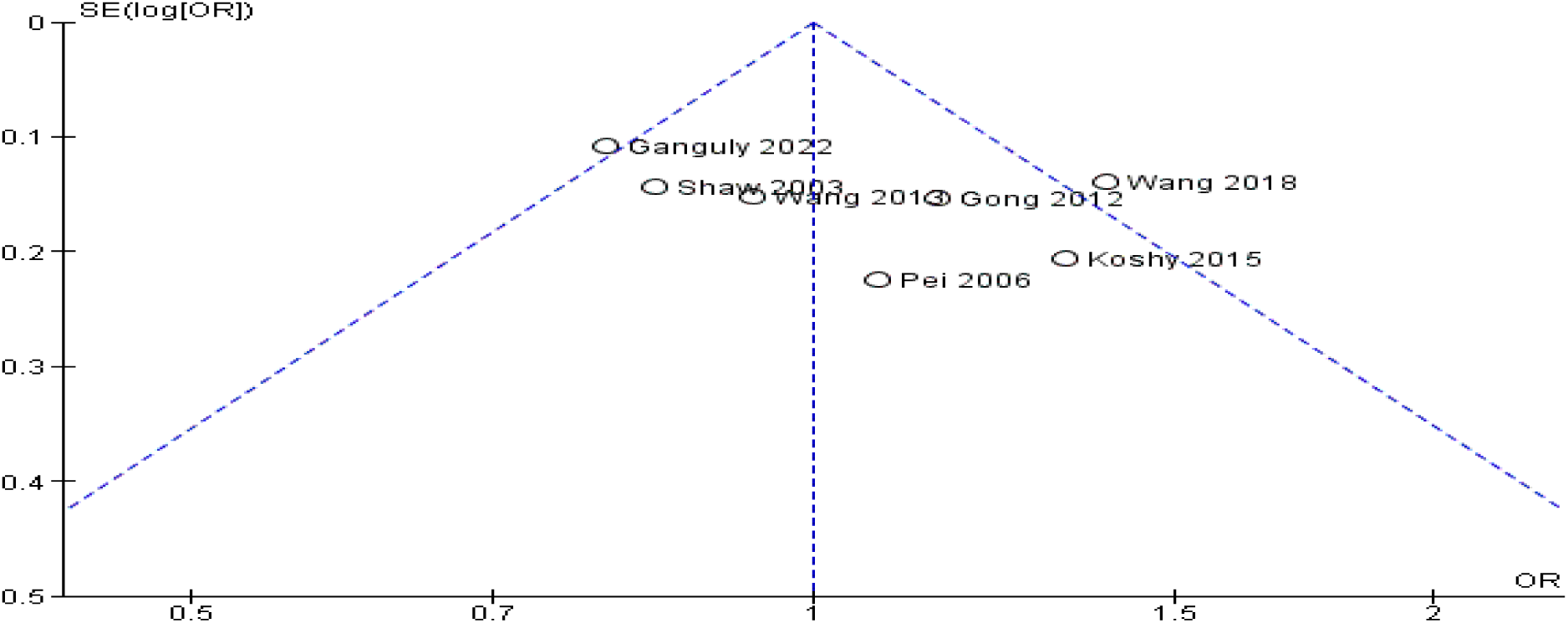
Funnel plot illustrating the identified outlier

**Figure 2.2.**
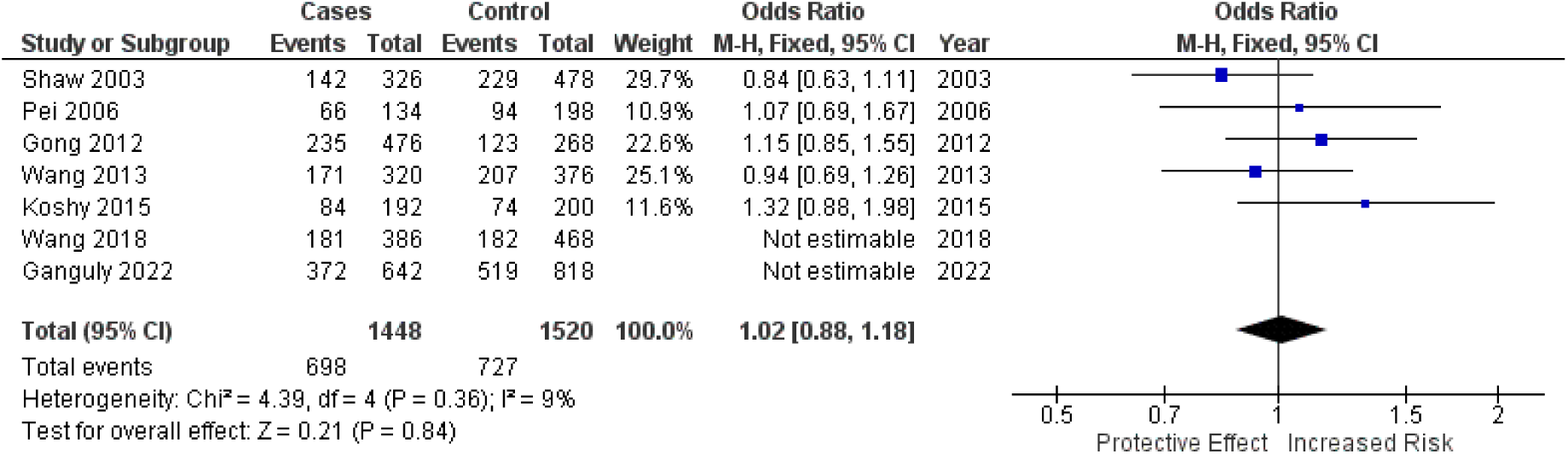
Forest plot following outlier removal

**Figure 3.**
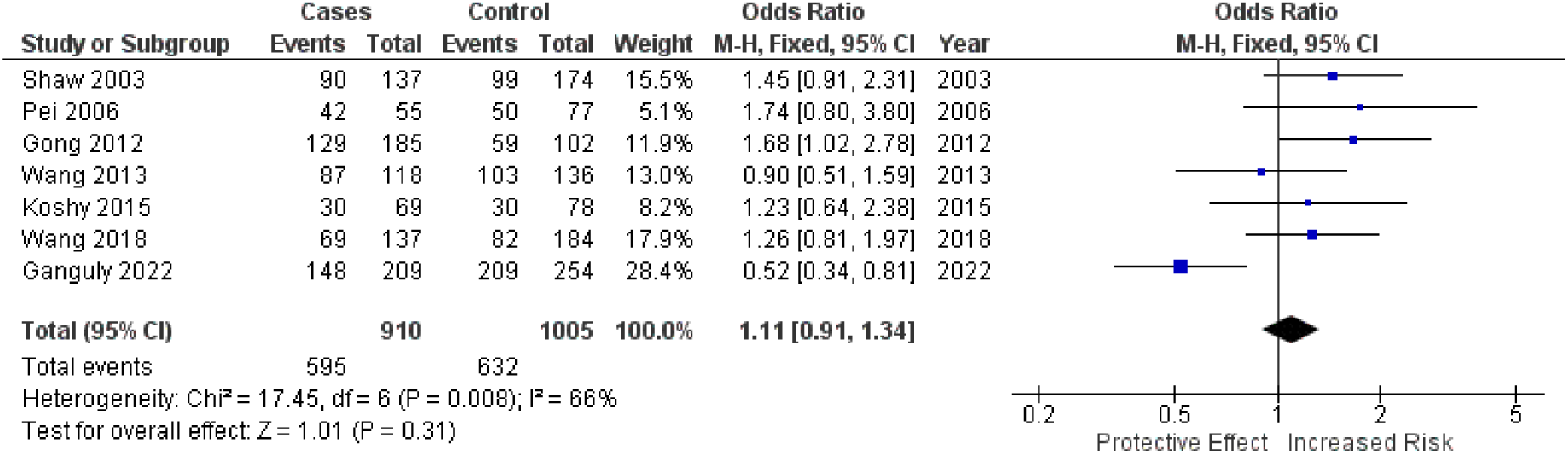
Forest plot prior to outlier removal

**Figure 3.1.**
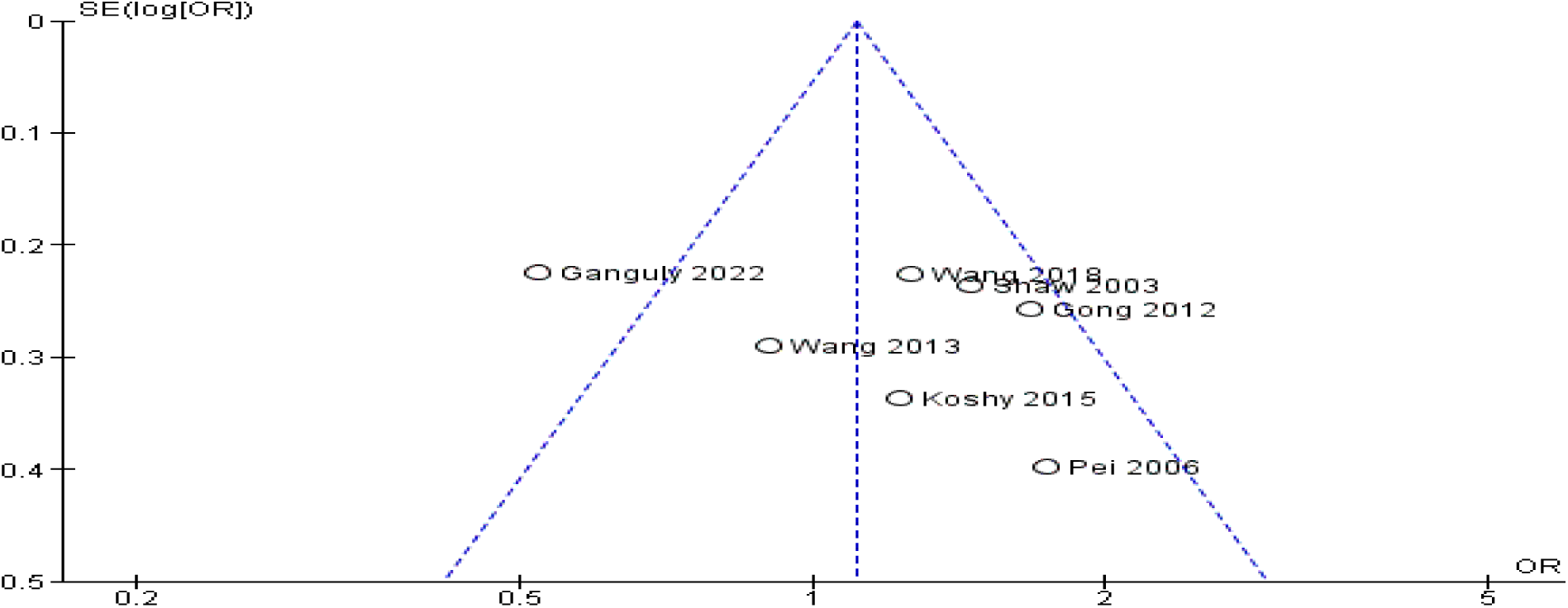
Funnel plot illustrating the identified outlier

**Figure 3.2.**
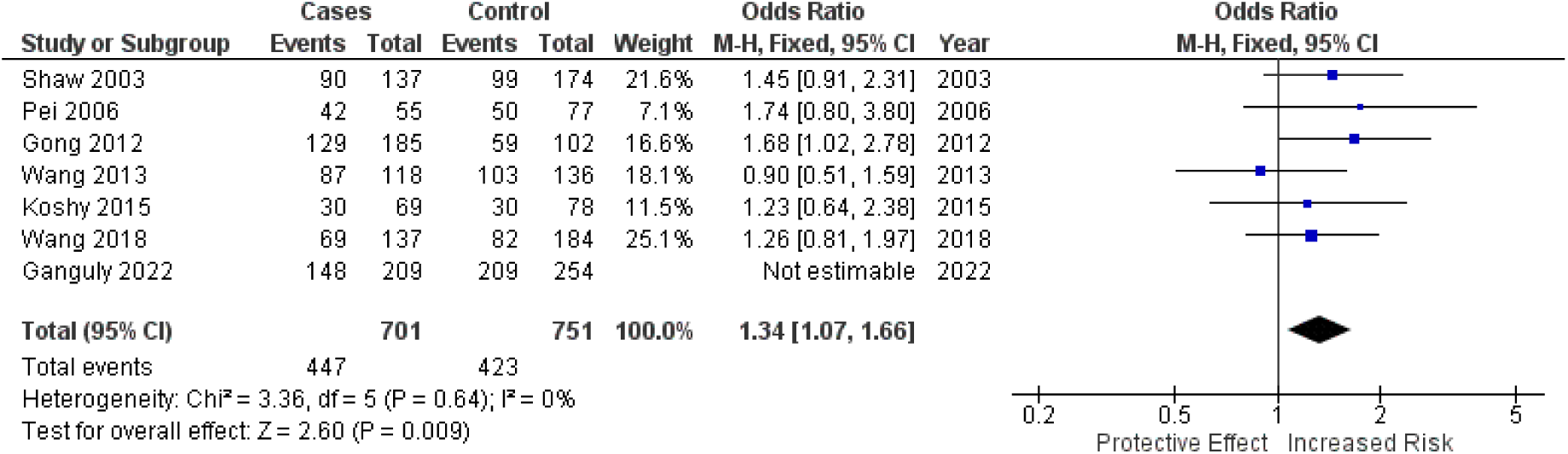
Forest plot following outlier removal

**Figure 4.**
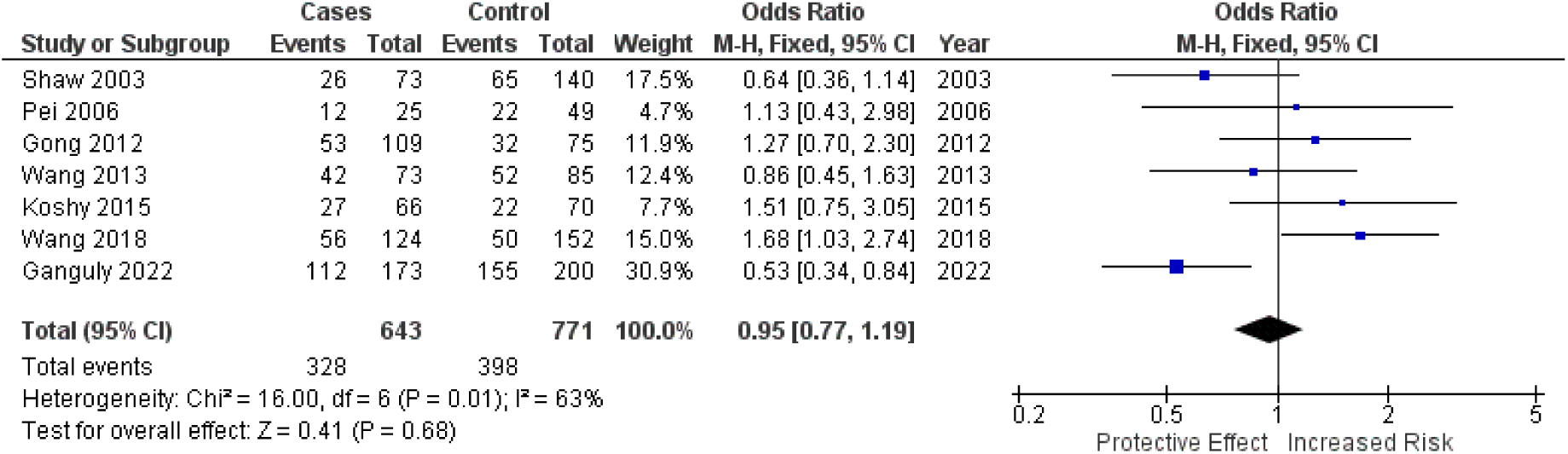
Forest plot prior to outlier removal

**Figure 4.1.**
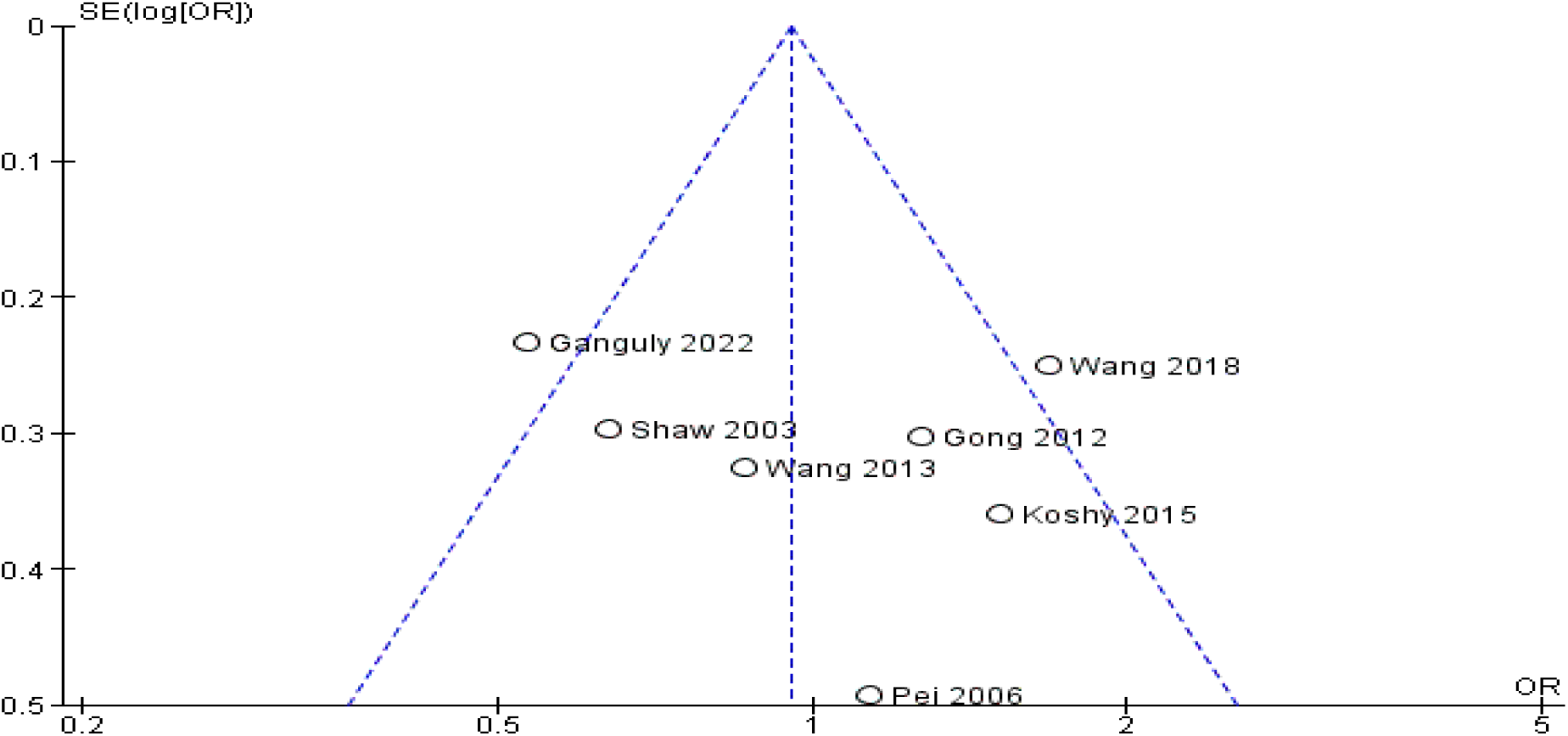
Funnel plot illustrating the identified outlier

**Figure 4.2.**
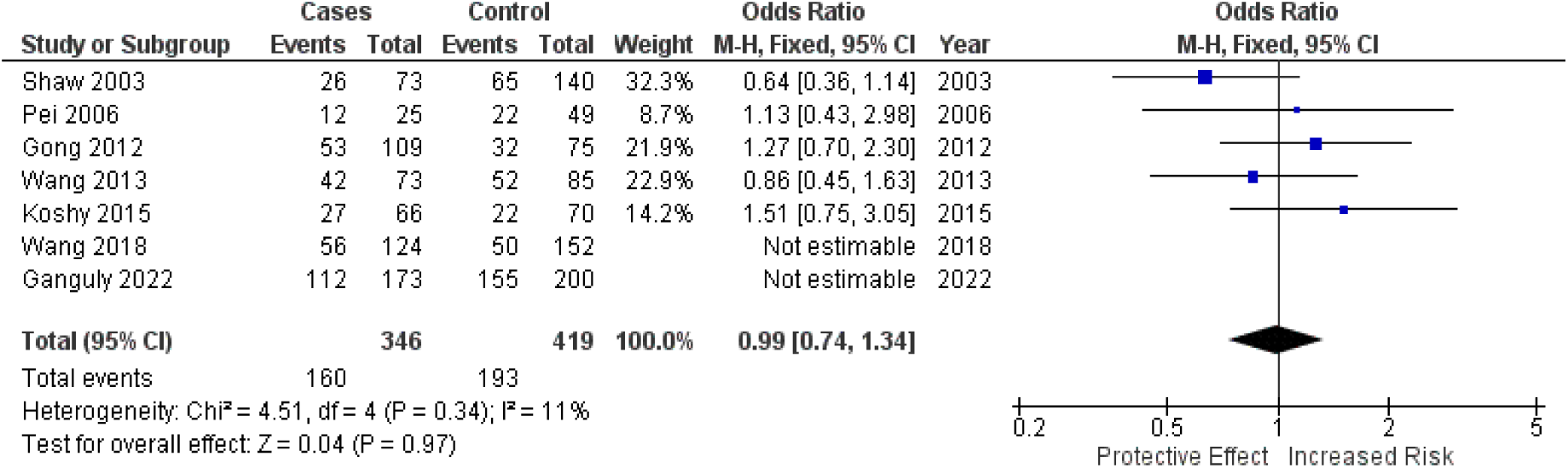
Forest plot following outlier removal

**Figure 5.**
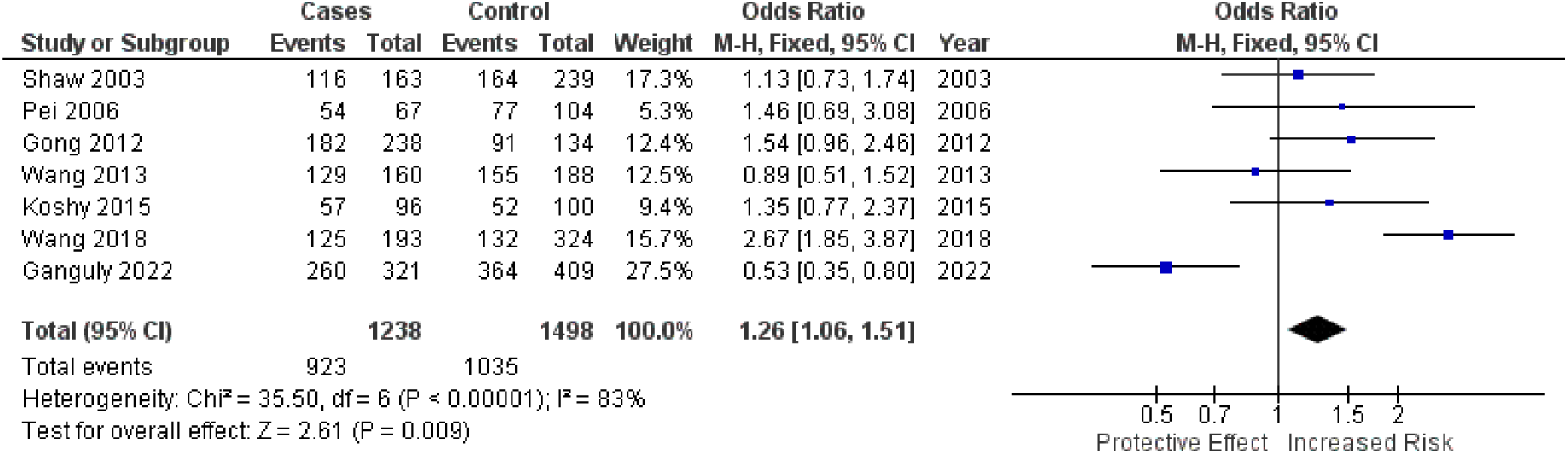
Forest plot prior to outlier removal

**Figure 5.1.**
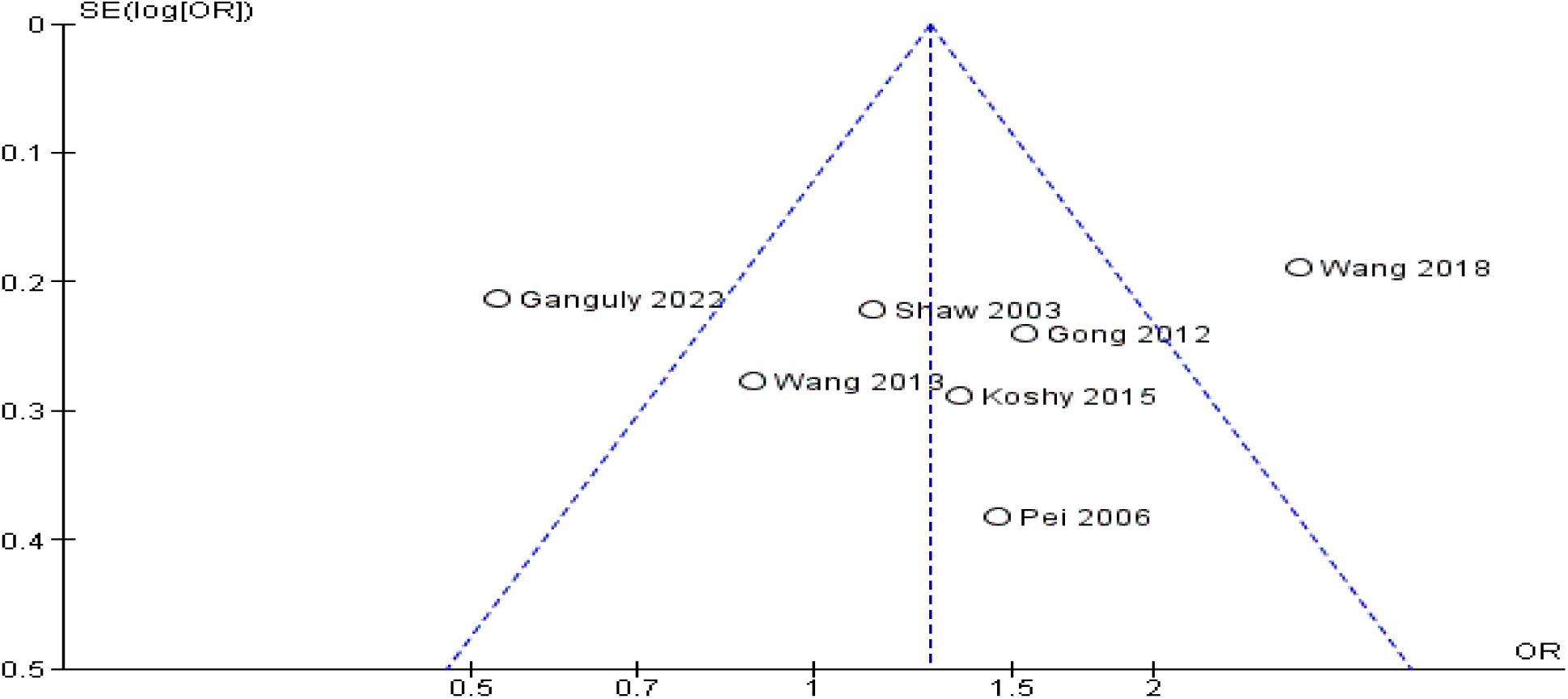
Funnel plot illustrating the identified outlier

**Figure 5.2.**
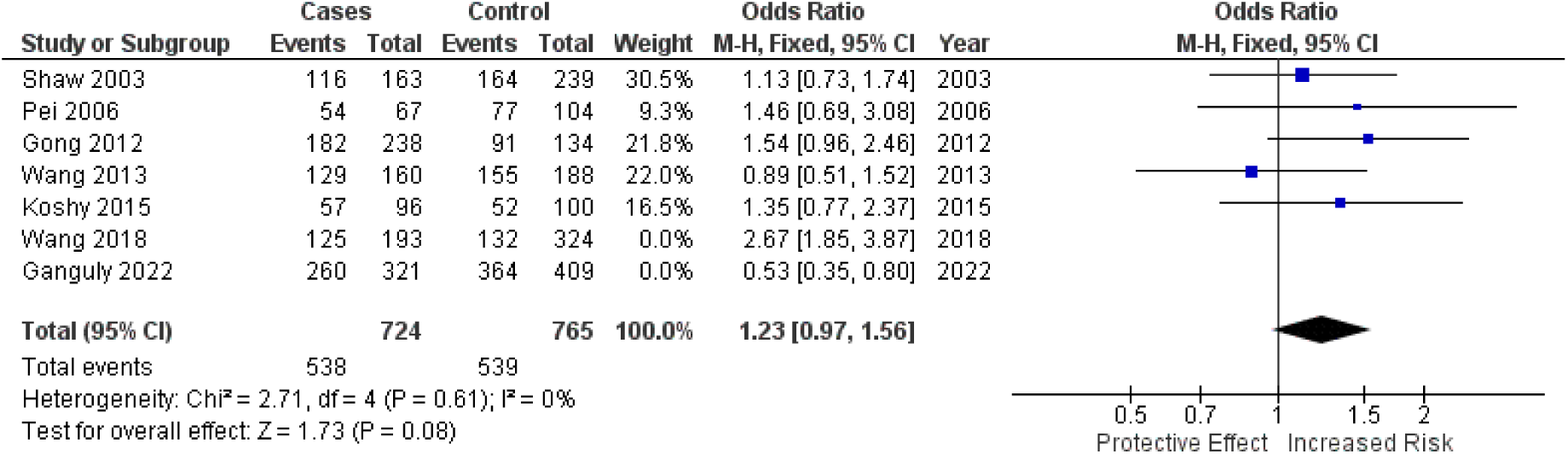
Forest plot following outlier removal

**Figure 6.**
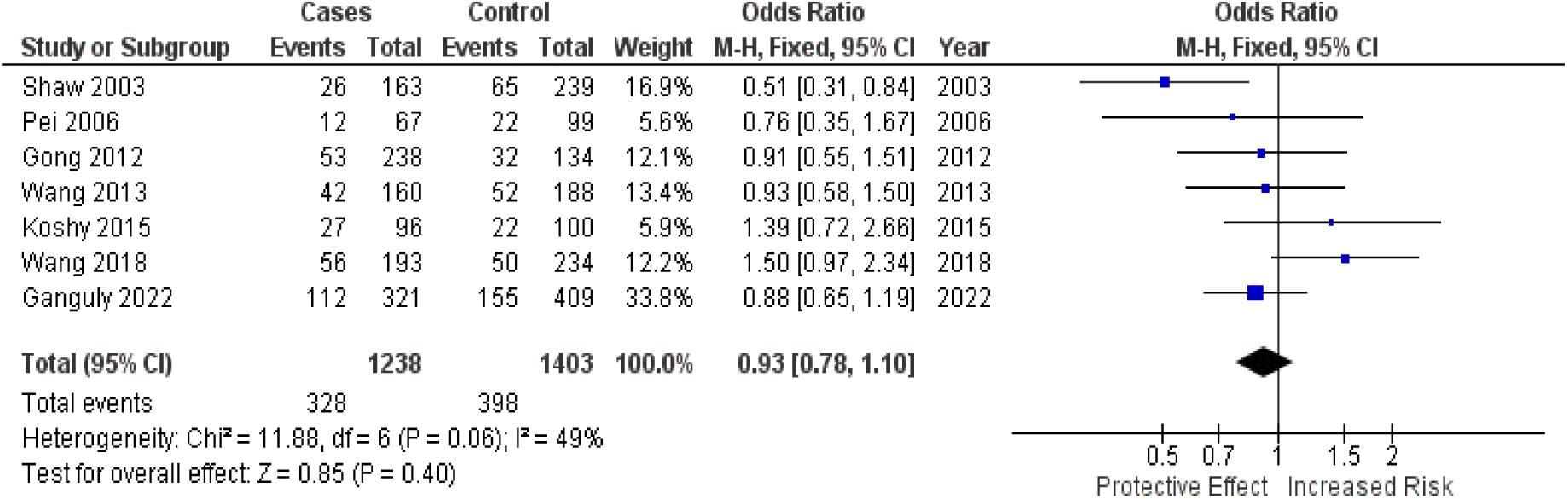
Forest plot prior to outlier removal

**Figure 6.1.**
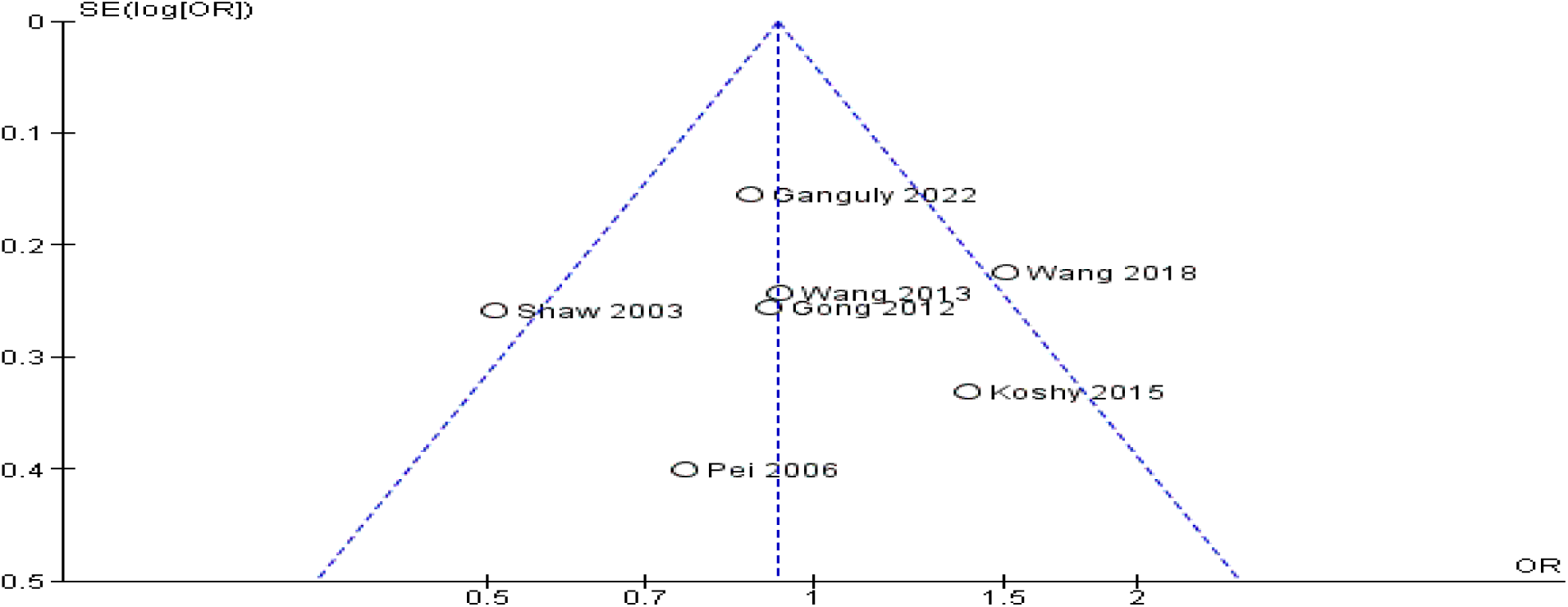
Funnel plot illustrating the identified outlier

**Figure 6.2.**
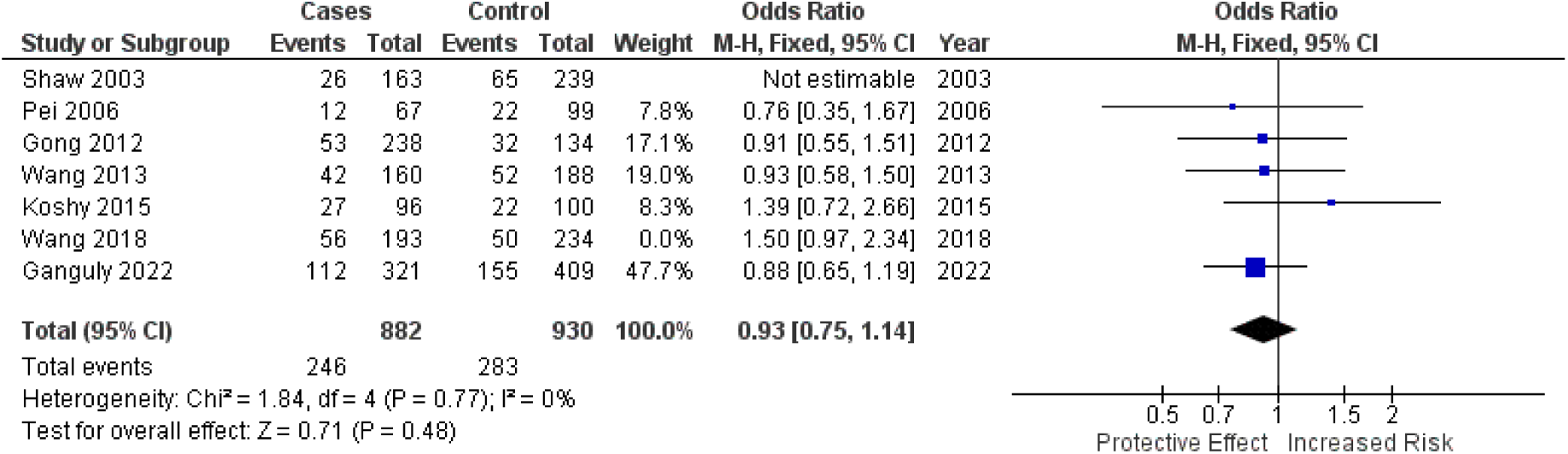
Forest plot following outlier removal

**Tabel 1.**

| Author/s | Year | Country | Region | Genotyping Method | Case Type | Control Source |
| --- | --- | --- | --- | --- | --- | --- |
| Wang et al. [13] | 2018 | China | East Asia | QIAamp DNA blood Mini kit | CTD | PB |
| Wang et al. [14] | 2013 | China | East Asia | SNaPShot multiplex PCR | CHD | HB |
| Gong et al. [15] | 2012 | China | East Asia | MALDI-ToF- Ms Method | TOG<br>TGA | HB |
| Shaw et al. [16] | 2003 | USA | North America | PCR- RFLP Analysis | CTD | PB |
| Koshy et al. [17] | 2015 | India | South Asia | Sanger Sequencing | CTD | HB |
| Pei et al. [18] | 2006 | China | East Asia | PCR-RFLP | CHD | PB |
| Ganguly et al. [20] | 2022 | India | South Asia | Bi-Directional Sanger sequencing<br>PCR | AVSD | HB |

**Keywords:** CTD - Conotruncal Heart Defects; CHD - Congenital Heart Defects; TOG - Tetralogy of Fallot; TGA - Transposition of the Great Arteries; AVSD - Atrioventricular Septal Defects; HB - Hospital based; PB - Population Based; MALDI ToF Ms - Matrix Assisted Laser Desorption/Ionization Time of Flight Mass Spectrometry; PCR - Polymerase Chain Reaction; RFLP - Restriction Fragment Length Polymorphism.

**Table 2.**
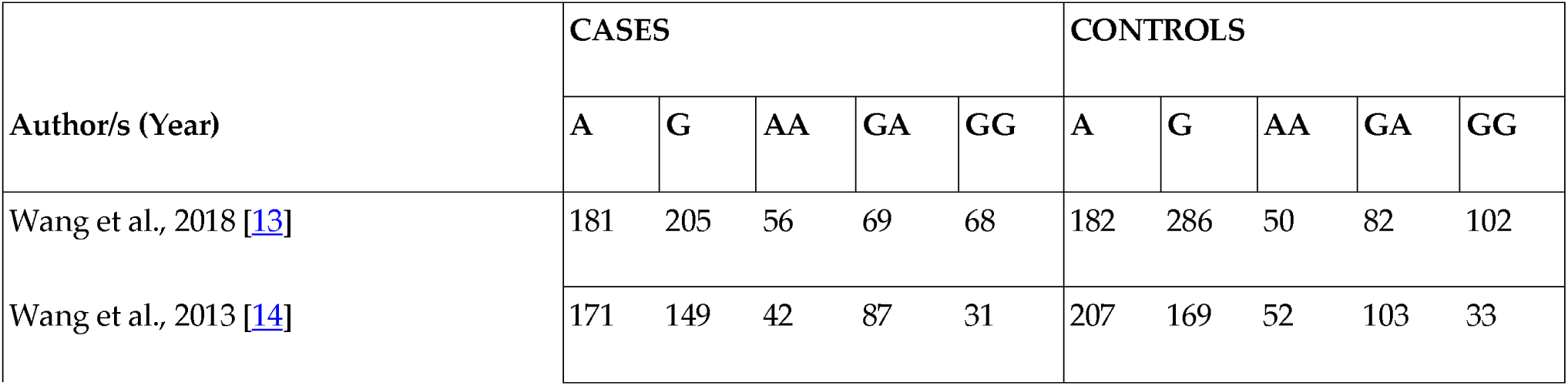
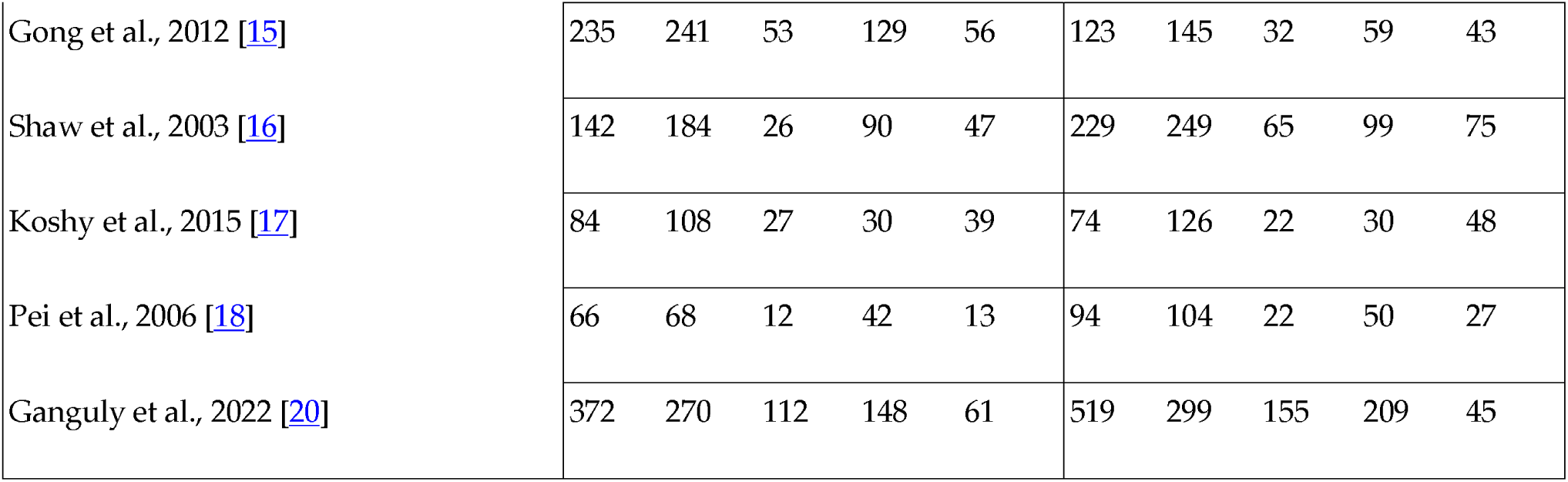
Genotypic Characteristics of Included Studies.

## Data Availability

All data produced in the present work are contained in the manuscript

## Author Contributions

Conceptualization, J.P.H; methodology, J.P.H, A.I.B, J.W.D.C, and K.G.S; software, J.P.H; validation, J.P.H, A.I.B, J.W.D.C, and K.G.S; formal analysis, , J.P.H, A.I.B, J.W.D.C, and K.G.S; investigation, , J.P.H, A.I.B, J.W.D.C, and K.G.S; resources, , J.P.H, A.I.B, J.W.D.C, and K.G.S; data curation, J.P.H; writing—original draft preparation, J.P.H; writing— review and editing, J.P.H, A.I.B, J.W.D.C, and K.G.S; visualization, J.P.H; supervision, J.P.H; project administration, J.W.D.C and K.G.S; All authors have read and agreed to the published version of the manuscript.

## Institutional Review Board Statement

The study protocol was registered to The International Prospective Register of Systematic Reviews (PROSPERO) under the UK’s National Institute for Health and Research (ID: CRD42024591902 and Date of Approval: 30/09/2024).

## Informed Consent Statement

Not applicable

## Data Availability Statement

The data supporting the findings of this study are not publicly available due to the complexities involved in gene extraction and processing. However, the extracted data and the raw files from the RevMan 5.4.1 analysis can be obtained from the corresponding author upon reasonable request.

## Acknowledgments

We sincerely thank Mr. Melandro Cunanan, M.Sc., and Ms. Lani Tabangay, M.Sc., for their invaluable guidance and support throughout this research. Their expertise and feedback have greatly influenced our understanding of the topic. We also appreciate the Angeles University Foundation (AUF) University Library for granting access to essential articles and research materials, which significantly aided our data gathering and literature review. The support from the library staff enhanced our research experience.

## Disclaimer/Publisher’s Note

The statements, opinions and data contained in all publications are solely those of the individual author(s) and contributor(s) and not of the editor(s). The editor(s) disclaim responsibility for any injury to people or property resulting from any ideas, methods, instructions or products referred to in the content.

